# Genetic liability to meniscus degeneration and its comorbidity patterns: a phenome-wide association study in the UK Biobank

**DOI:** 10.1101/2025.09.23.25336468

**Authors:** Dong-gi Lee, Yonghyun Nam, Jaehyun Joo, Se-Hwan Lee, Jaeun Jung, Su Chin Heo, Dokyoon Kim

## Abstract

Meniscus degeneration is a common knee pathology causing pain, disability, and osteoarthritis. While mechanical factors are established, the contribution of inherited genetic liability to its systemic disease patterns remains unclear. We applied a polygenic risk score (PRS) for meniscus degeneration, derived from FinnGen genome-wide association study (GWAS) results to 323,999 UK Biobank participants and conducted a phenome-wide association study (PRS-PheWAS) across 962 clinical phenotypes. The PRS-PheWAS revealed significant associations beyond musculoskeletal traits, extending to metabolic, cardiovascular, psychiatric, and gastrointestinal domains, indicating broad shared genetic architecture. To support these findings, linkage disequilibrium score regression confirmed strong genetic correlations with osteoarthritis and related arthropathies, and genotype-tissue expression (GTEx) analysis highlighted tissue-specific expression of lead genes (e.g., GDF5, SOX5, BMP6) in connective and metabolic tissues. The results demonstrate that genetic liability to meniscus degeneration extends beyond the knee, sharing pathways with systemic conditions. This systemic genetic architecture underscores the need for integrative management approaches that combine orthopedic care with metabolic and lifestyle interventions.

## 1. Introduction

Meniscus degeneration is a common intra-articular knee pathology that contributes to chronic knee pain and functional limitation, often preceding the development of knee osteoarthritis (OA)^1, 2^. Degenerative meniscal tears typically occur in middle-aged and older adults without a specific acute injury, and they are strongly linked to subsequent radiographic and symptomatic knee OA^3^. Traditional risk factors for meniscal degeneration include age-related tissue wear, occupational or sports-related repetitive knee trauma, obesity-related excess mechanical loading, and prior knee injuries such as anterior cruciate ligament ruptures^2, 4^. However, these well-recognized mechanical and environmental contributors do not fully explain the substantial inter-individual variability in meniscus degeneration onset and progression^1, 5, 6^. The clinical impact of meniscus degeneration is considerable. Once symptomatic, patients often experience persistent pain and functional limitation, with an elevated risk of progressing to knee OA^2^. Despite its prevalence, no FDA-approved disease-modifying therapies currently exist^7^. Management is largely restricted to symptom alleviation, including physical therapy, lifestyle modification, and intra-articular injections, while advanced cases often necessitate surgical interventions such as partial meniscectomy or meniscal repair^8, 9^. These limitations highlight the urgent need to better define disease etiology and risk architecture in order to guide preventive and therapeutic strategies.

Emerging evidence indicates that genetic predisposition and systemic comorbidities may also play important roles in determining susceptibility to meniscal pathology. Twin studies indicate that meniscal tears have a moderate genetic heritability of ∼40%, underscoring that inherited factors significantly influence susceptibility^10^. In addition, meniscal pathology often coexists with systemic conditions (e.g., obesity and metabolic disorders), suggesting that factors beyond localized joint stress may play an etiologic role. Recent evidence from a Mendelian randomization analysis has demonstrated a causal link between a higher body mass index and an increased risk of meniscal injury (odds ratio ∼1.13 per unit increase in body mass index), reinforcing the contribution of metabolic factors to meniscus degeneration^11^. These findings motivate a deeper exploration of the genetic and systemic context in which meniscal degeneration occurs. In addition, patients with chronic joint conditions also frequently experience depression, anxiety, and other mental health challenges, pointing to a psychosocial dimension in the disease trajectory^12^. Yet, few studies have systematically mapped out the full spectrum of comorbidities and shared risk factors that accompany meniscal degeneration. Prior investigations have typically focused on one risk factor or comorbidity at a time (e.g., obesity or a single metabolic marker), leaving a gap in understanding how multiple coexisting conditions collectively relate to meniscus health. Identifying the broader phenotypic network associated with meniscus degeneration could yield insights into underlying pathogenic mechanisms. For example, it reveals whether meniscal degeneration shares a genetic architecture with systemic metabolic dysregulation, inflammation, or neuropsychiatric factors. This knowledge may help stratify individuals into meaningful risk endotypes and suggest interdisciplinary intervention strategies.

In this context, a polygenic risk score (PRS), which aggregates the effects of numerous genetic variants associated with a trait, offers a framework to investigate whether genetic liability to meniscus degeneration is shared across diverse phenotypes^13^. Phenome-wide association studies (PheWAS) of PRS have been used to identify broad genotype-phenotype associations, revealing how polygenic liability to one trait relates to diverse disease outcomes^14^. We hypothesize that applying PRS-PheWAS to meniscus degeneration will reveal its comorbidity landscape, defined by diseases that share underlying genetic risk.

In this study, we applied a PRS-based phenome-wide association study (PRS-PheWAS) in the UK Biobank (UKB) to identify comorbidity patterns associated with genetic liability to meniscus degeneration. This framework provides a system-level perspective to inform integrated strategies for prevention and management beyond the knee joint itself.

## 2. Results

### 2.1. Study overview and cohort characteristics

We investigated the genetic risk architecture of meniscus degeneration by constructing a polygenic risk score (PRS) from FinnGen GWAS summary statistics and applying it to 323,999 participants of the UKB, enabling a phenome-wide assessment of associations with genetic liability across 962 disease phenotypes (**Fig. 1**). We validated the PRS against meniscal pathology within UKB, characterized its associations across musculoskeletal and systemic traits, and quantified genetic correlations with related conditions using LD score regression. Finally, we integrated transcriptomic data from GTEx to contextualize lead GWAS genes by tissue-specific expression.

**Figure 1.**
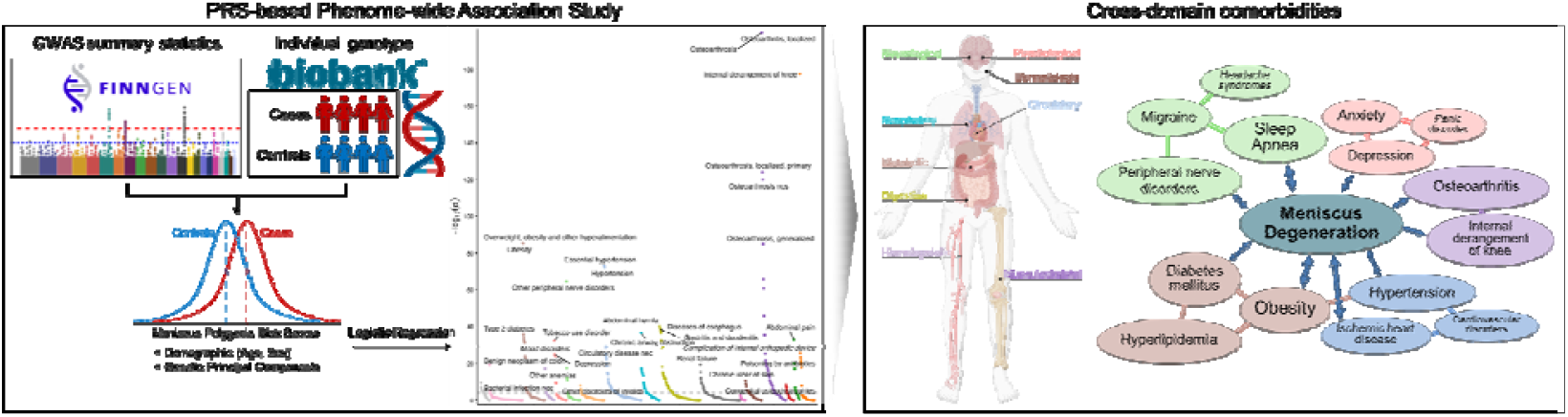
Overview of study design.

We analyzed 323,999 individuals with high genetic similarity with European reference populations from the UKB (see **Methods**). The mean baseline age was 56.9 years, and 46.5% were male. Meniscus degeneration was identified in 11,829 individuals (3.7%). Additional cohort characteristics are summarized in **Table 1**.

**Table 1.** Cohort characteristics of participants by polygenic risk score quartiles

|  | All | PRS Quartile Groups |  |  |  | P* |
| --- | --- | --- | --- | --- | --- | --- |
|  |  | PRS Q1 | PRS Q2 | PRS Q3 | PRS Q4 |  |
| N | 323,999 | 81,000 | 81,000 | 81,001 | 80,998 |  |
| Meniscus degeneration % | 11,829 (3.7) | 2,090 (2.6) | 2,636 (3.3) | 3,166 (3.9) | 3,937 (4.9) | < 0.001 |
| Age, mean (sd) | 56.91 (7.96) | 56.98 (7.96) | 56.91 (7.96) | 56.82 (7.99) | 56.93 (7.94) | 0.012 |
| Male, N (%) | 150,500 (46.5) | 37,608 (46.4) | 37,483 (46.3) | 37,744 (46.6) | 37,715 (46.6) | 0.174 |
| Ever smoker, N (%) |  |  |  |  |  | < 0.001 |
| Yes | 194,790 (60.1) | 48,234 (59.5) | 48,421 (59.8) | 48,959 (60.4) | 49,176 (60.7) |  |
| No | 128,126 (39.5) | 32,526 (40.2) | 32,294 (39.9) | 31,796 (39.2) | 31,537 (38.9) |  |
| Not available | 1,083 (0.3) | 240 (0.3) | 285 (0.4) | 273 (0.3) | 285 (0.4) |  |
| BMI†, N (%) |  |  |  |  |  | < 0.001 |
| Underweight | 1,619 (0.5) | 477 (0.6) | 429 (0.5) | 366 (0.5) | 347 (0.4) |  |
| Normal weight | 105,433 (32.5) | 28,875 (35.6) | 26,933 (33.3) | 25,699 (31.7) | 23,956 (29.6) |  |
| Overweight | 137,994 (42.6) | 34,396 (42.5) | 34,395 (42.5) | 34,699 (42.8) | 34,504 (42.6) |  |
| Obesity | 77,906 (24.0) | 17,027 (21.0) | 18,970 (23.4) | 19,987 (24.7) | 21,922 (27.1) |  |
| Not available | 1,047 (0.3) | 225 (0.3) | 273 (0.3) | 280 (0.3) | 269 (0.3) |  |

\*P values are based on univariate tests using the continuous PRS values; †BMI categories: underweight (BMI < 18.5 kg/m<sup>2</sup>), normal weight (18.5 ≤ BMI < 25), overweight (25 ≤ BMI < 30) and Obesity (BMI ≥ 30).

To establish the validity of the PRS prior to phenome-wide analyses, we examined its distribution and association with meniscal pathology within UKB. Demographic characteristics were generally well balanced across PRS quartiles, with comparable mean age and sex distribution. However, meniscus degeneration prevalence increased steadily with higher PRS quartiles, from 2.6% in Q1 to 4.9% in Q4 (P < 0.001). Smoking history also showed a slight upward trend in higher quartiles (P < 0.001). Body mass index (BMI) distributions differed significantly across quartiles, with lower proportions of normal weight and higher proportions of obesity in the top PRS group (27.1% in Q4 vs. 21.0% in Q1; P < 0.001). A one-standard-deviation increase in PRS was associated with a significantly elevated risk of meniscu degeneration (OR = 1.31; 95% CI = 1.29–1.33; P < 0.001). This positive control confirms that the PRS captures meaningful genetic liability for meniscus degeneration in the UKB cohort and provides a basis for mapping its association with other health outcomes across the phenome.

### 2.2. Phenome-wide associations of meniscus degeneration genetic risk

Using a PRS-PheWAS framework, we evaluated 962 clinical phenotypes and identified 216 significant associations at the Bonferroni threshold (P < 5.20×10^-5^)(**Fig. 2a**; **Supplementary Table S1**). Consistent with the target phenotype, the musculoskeletal domain yielded the strongest signals, with highly significant associations for osteoarthrosis and joint pain, providing internal validation of the PRS approach. Importantly, associations extended beyond musculoskeletal traits to include metabolic, cardiovascular, and gastrointestinal domains, highlighting broader patterns of disease linked to genetic liability for meniscus degeneration. At the phenotype level, osteoarthrosis (P < 1×10^-300^), internal derangement of the knee (P = 3.75×10^-178^), and joint pain (P = 9.84×10^-62^) were among the most significant findings, the latter serving as an expected positive control given its direct clinical relevance. Associations with abdominal pain (P = 9.61×10^-34^), and back pain (P = 7.49×10^-21^) suggest that genetic risk may manifest as broader pain-related conditions across the phenome.

**Figure 2.**
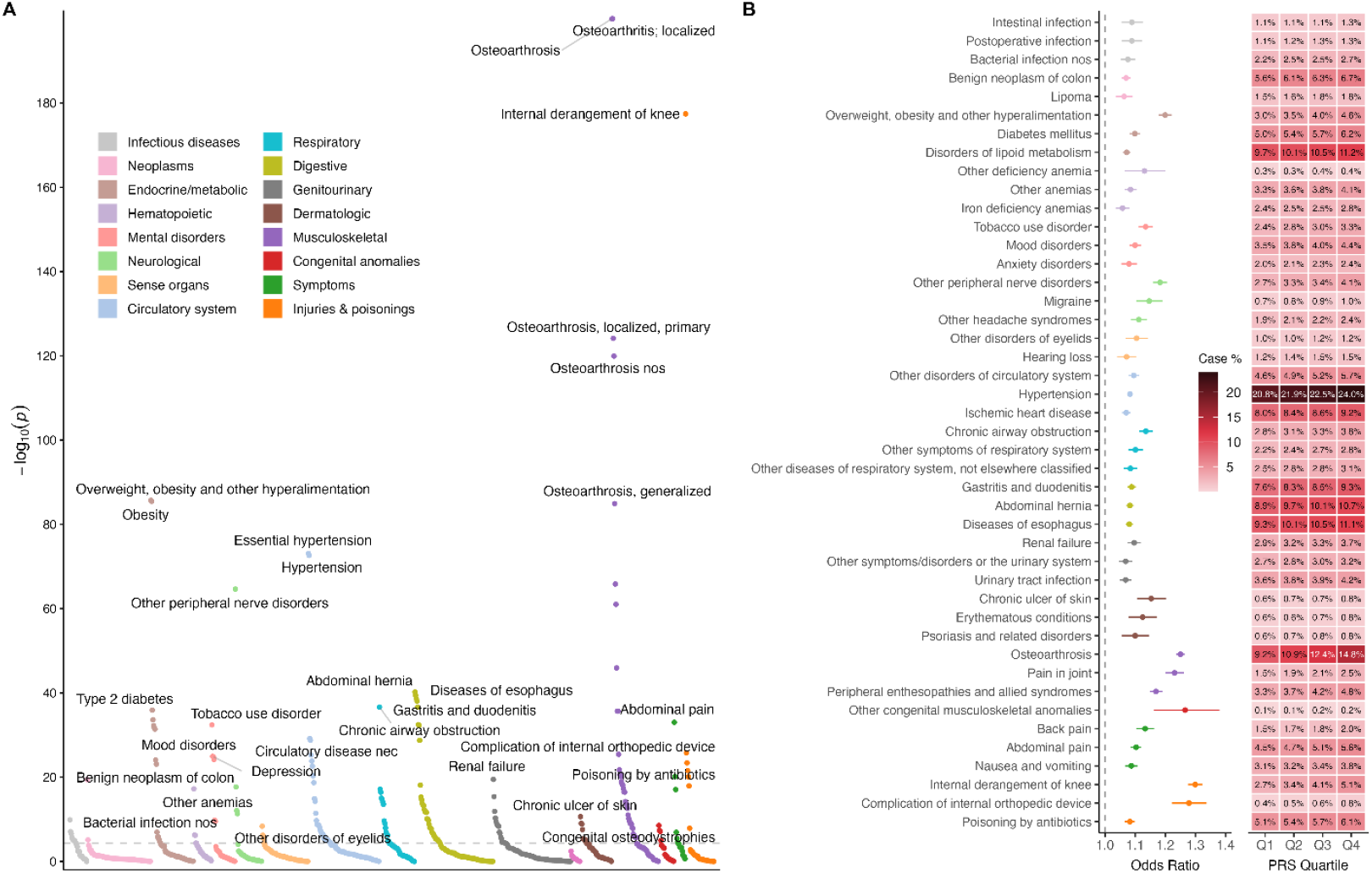
Phenome-wide association study (PheWAS) of a polygenic risk score for meniscus degeneration. A) The Manhattan plot shows the statistical significance of associations between the genetic risk score and a wide range of phenotypes. The y-axis represents the significance, and the dashed horizontal line indicates the Bonferroni-corrected threshold for statistical significance. Phenotypes, represented as individual points, are grouped by physiological system on the x-axis. B) The forest plot illustrates the effect sizes (odds ratios, ORs) and their 95% confidence intervals (CIs) for the top three most significant root phenotypes within each disease category that reached Bonferroni-corrected significance. An OR greater than 1.0 indicates increased odds of the disease per standard deviation increase in the polygenic risk score. The heatmap displays the disease prevalence (case %) across the four quartiles (Q1–Q4) of the polygenic risk score. The color intensity corresponds to the percentage of cases in each quartile.

Notably, the meniscus degeneration PRS was linked to a wide range of systemic phenotypes in addition to musculoskeletal outcomes (**Fig. 2b**). In the metabolic domain, overweight/obesity, type 2 diabetes, and lipid disorders showed strong signals. Cardiovascular phenotypes included hypertension and ischemic heart disease. In the mental health domain, depression, anxiety, and tobacco use disorder were significantly associated. Neurological outcomes such as peripheral nerve disorders and migraine, respiratory conditions including chronic airway obstruction, and gastrointestinal phenotypes such as gastroesophageal reflux disease, gastritis/duodenitis, and abdominal hernia also reached significance. Additional associations were observed in dermatologic (chronic ulcer of skin, psoriasis), genitourinary (renal failure, urinary tract infection), and infectious disease categories. These findings demonstrate that the meniscus PRS not only recapitulates known musculoskeletal links but also points to novel cross-domain associations, illustrating its utility for mapping potential comorbidity patterns at the phenome-wide scale.

### 2.3. Sensitivity analysis

#### Exclusion PheWAS

We evaluated whether the phenome-wide associations of the meniscus degeneration PRS were driven by the presence of individuals with a clinical diagnosis of meniscus degeneration. In sensitivity analyses adjusting for meniscus degeneration status as a covariate and excluding all diagnosed cases, the overall pattern and magnitude of associations were highly consistent with the primary findings (**Fig. 3A**; **Table 2**). Consistent with the target phenotype, the phenotypes directly related to meniscus degeneration, such as internal derangement of knee within injuries & poisoning category, showed the largest effect size in the baseline model (OR = 1.30; P = 2.2×10□^1^ □). This association was attenuated but remained nominally significant when adjusting for meniscus degeneration status (OR = 1.12; P=7.81×10 □ □) and in the case-excluded analysis (OR = 1.17; P = 4.58×10 □ □), suggesting that the PRS captures overlapping genetic risk across meniscus-related condition. These results indicate that the observed PheWAS associations are not solely explained by prevalent meniscus degeneration cases but reflect broader shared genetic influences of the meniscus risk across diverse disease domains.

**Table 2.**
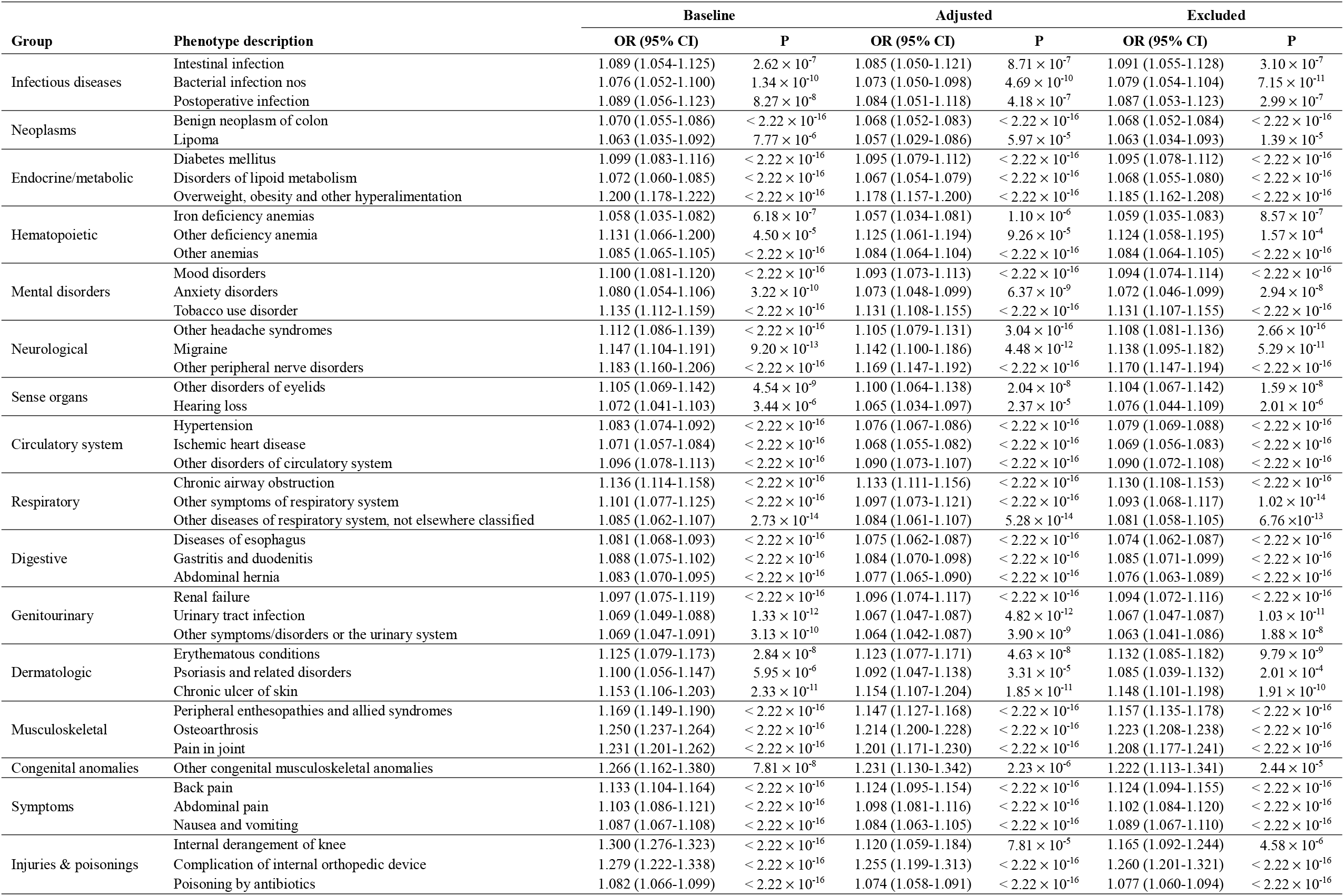
Sensitivity analysis of meniscus degeneration PRS with significant associations in the UK Biobank

**Figure 3.**
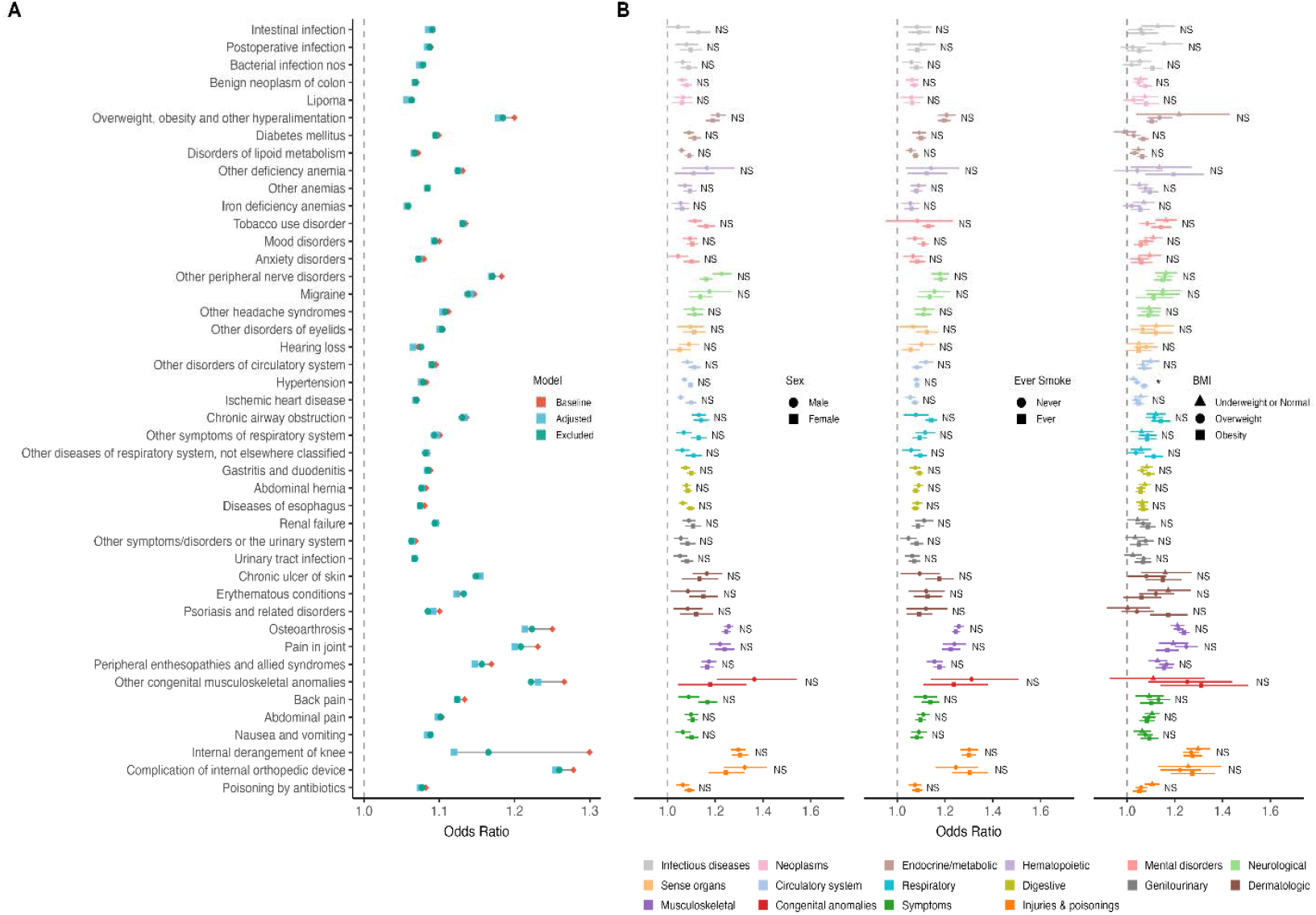
Sensitivity analyses of meniscus degeneration polygenic risk score (PRS) with phenotypes in the UK Biobank. A) The dumbbell plot shows odds ratios (ORs) with PRS and covariates (baseline), meniscus degeneration status as a covariate (adjusted), and excluding all diagnosed cases (excluded). B) Forest plot for stratified associations. ORs with 95% confidence intervals are shown for selected root-level phenotypes, stratified by sex (left), smoking history (center), and BMI category (right). Phenotypes are color-coded by disease categories. Statistical significance from interaction tests with Bonferroni correction, which assess whether the effect of the PRS on a given outcome differs between subgroups, is indicated as *P* < 0.05 (*), NS for non-significant results.

#### Sex, smoking status, and BMI stratified analyses

To further examine the robustness of these associations, we assessed stratified analysis whether the effects of the PRS varied by demographic or lifestyle factors, including sex, smoking status, and BMI (**Fig. 3B**). Overall, the associations of the PRS were consistent across subgroups, and most interaction tests were not statistically significant. A significant interaction was observed between the PRS and BMI for hypertension; the effect of the PRS was more pronounced at higher BMI, with the largest odds ratios seen in the obese subgroup. No significant interactions were detected by sex and smoking status; notably, the association with tobacco use disorders was present only among ever smokers and absent among never smokers.

### 2.4. Genome-wide genetic correlations of meniscus degeneration

To complement the PRS-PheWAS findings, we assessed genome-wide genetic correlations (r_g_) between meniscus degeneration and a broad range of traits using LD score regression with FinnGen GWAS summary statistics. Strong positive correlations were observed with multiple musculoskeletal phenotypes (**Supplementary Table S2**). The largest signals were observed for knee arthrosis and related arthropathies (r_g_ = 0.88), arthrosis at multiple sites (r_g_=0.82), and joint pain (r_g_=0.68). Substantial correlations were also evident for enthesopathies, synovitis/tenosynovitis, and soft tissue overuse injuries (r_g_=0.66–0.70). Beyond joint-specific disorders, meniscus degeneration demonstrated strong genetic correlations with plantar fasciitis (r_g_=0.81) and brachial plexus disorders (r_g_=0.71), suggesting broader shared polygenic architecture across connective tissue and peripheral nerve-related conditions. These results are consistent with and extend the PRS-PheWAS associations, reinforcing the conclusion that genetic risk for meniscus degeneration overlaps with polygenic influences on musculoskeletal and related disorders.

### 2.5. Functional characterization of meniscus-associated genes using GTEx

The PRS-PheWAS revealed associations of genetic risk for meniscus degeneration not only with musculoskeletal conditions but also with diverse non-musculoskeletal domains, while LDSC analysis demonstrated polygenic correlations with related musculoskeletal phenotypes. While these statistical approaches delineate genetic associations, they do not clarify whether implicated loci operate through specific biological pathways or tissues. As a complementary analysis, we examined tissue-level expression profiles of genome-wide significant genes from the FinnGen GWAS (used for PRS construction) using GTEx v10 median transcript abundance, to contextualize association signals within relevant biological systems.

Nine lead genes were prioritized, including GDF5, SMG6, DST, LYPLAL1-AS1, SOX5, CFAP299, BMP6, TACC3, and DLEU1. Several exhibited tissue-specific expression patterns (**Fig. 4**; **Table 3**; complete list is available in **Supplementary Table S3**). SOX5 and GDF5 were enriched in fibroblast and connective tissue-related contexts, consistent with roles in skeletal development and joint morphogenesis. BMP6 showed preferential expression in adipose tissue, aligning with PRS-PheWAS associations with obesity and metabolic traits. In contrast, DST and SMG6 exhibited broader profiles, with the highest levels in skin and ovary, suggesting systemic roles beyond musculoskeletal tissues. Other genes, including CFAP299, TACC3, and DLEU1, displayed strong testis-specific signals.

**Table 3.**
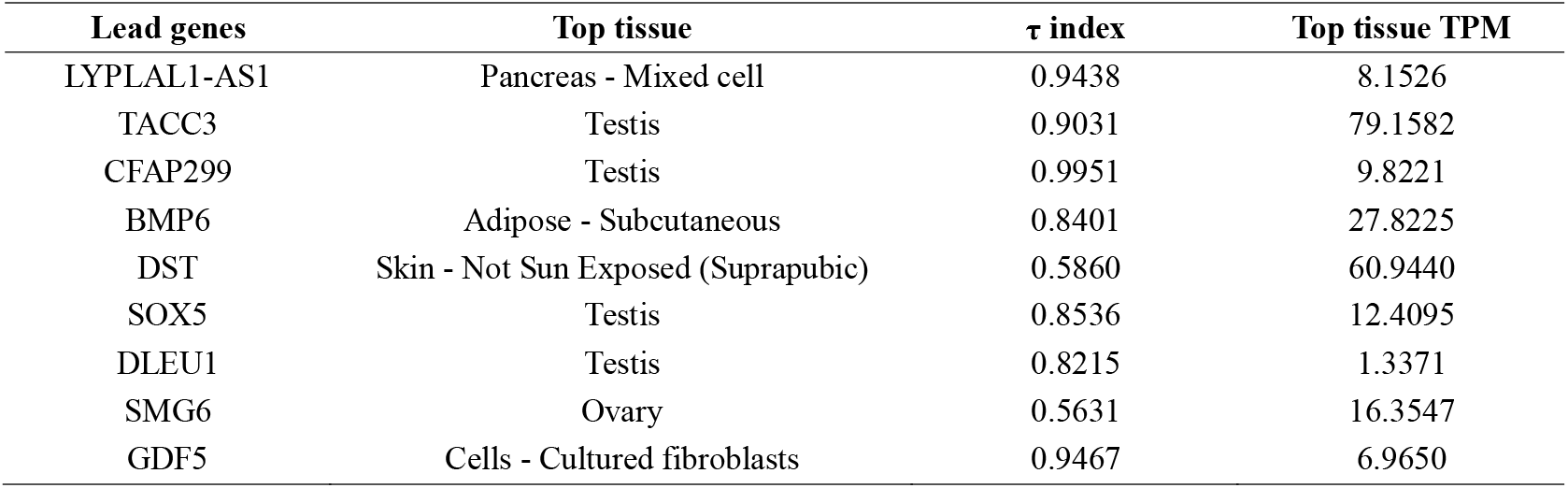
Tissue specificity of meniscus degeneration lead genes. τ index quantifies tissue specificity (0 = broad expression; 1 = tissue-restricted). The top tissue and its median expression level are reported for <u>each gene.</u>

| Lead genes | Top tissue | $\tau$ index | Top tissue TPM |
| --- | --- | --- | --- |
| LYPLAL1-AS1 | Pancreas - Mixed cell | 0.9438 | 8.1526 |
| TACC3 | Testis | 0.9031 | 79.1582 |
| CFAP299 | Testis | 0.9951 | 9.8221 |
| BMP6 | Adipose - Subcutaneous | 0.8401 | 27.8225 |
| DST | Skin - Not Sun Exposed (Suprapubic) | 0.5860 | 60.9440 |
| SOX5 | Testis | 0.8536 | 12.4095 |
| DLEU1 | Testis | 0.8215 | 1.3371 |
| SMG6 | Ovary | 0.5631 | 16.3547 |
| GDF5 | Cells - Cultured fibroblasts | 0.9467 | 6.9650 |

**Figure 4.**
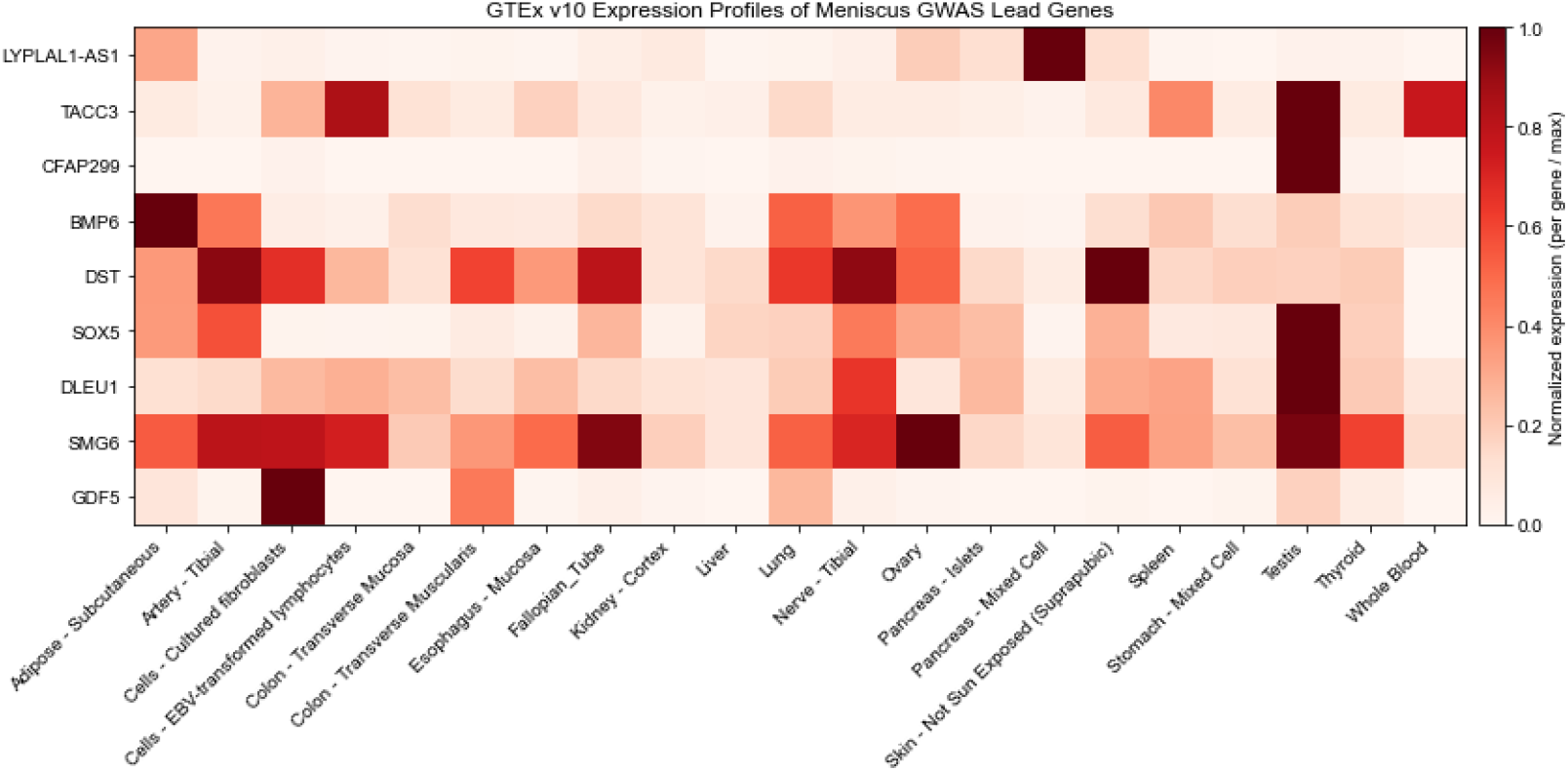
GTEx tissue expression profiles of lead genes associated with meniscus degeneration. Heatmap shows median transcript per million (TPM) values across GTEx v10 tissues, highlighting tissue-specific and broadly expressed patterns.

The tissue-specificity index (τ) further highlighted these distinctions. CFAP299 demonstrated the highest specificity (τ = 0.99), followed by GDF5 (τ = 0.95), LYPLAL1-AS1 (τ = 0.94), and SOX5 (τ = 0.85). Genes with lower τ indices, such as DST (τ = 0.59) and SMG6 (τ = 0.56), were broadly expressed across tissues. These results indicate that meniscus-associated loci include both tissue-specific regulators of skeletal and metabolic pathways as well as broadly expressed genes implicated in systemic processes.

## 3. Discussion

In this PRS-PheWAS study, we found that polygenic risk for meniscus degeneration extends well beyond localized knee pathology, showing associations with musculoskeletal, metabolic, cardiovascular, psychiatric, and inflammatory conditions. These findings suggest that meniscus degeneration, traditionally viewed as a focal orthopedic disorder, shares systemic genetic influences with a wide spectrum of diseases. This aligns with emerging concepts in osteoarthritis research that emphasize the interplay of metabolic imbalance, chronic inflammation, and neurosensory modulation of pain in joint^15^.

Among the most consistent signals was overlap with metabolic traits, including obesity, adverse lipid profiles, hypertension, and type 2 diabetes. These associations reinforce clinical observations that obesity is a major risk factor for degenerative meniscal tears11 and support the concept of “metabolic osteoarthropathy^16-18^. Beyond mechanical loading, adiposity-related inflammatory mediators may represent a shared biological mechanism linking systemic metabolic dysregulation with cartilage and meniscal degeneration. Psychiatric and behavioral traits, including depression, anxiety, and tobacco use disorder, were also enriched, suggesting shared polygenic influences on pain sensitivity, mood regulation, and addictive behaviors^19-21^. Associations with autoimmune and inflammatory traits such as psoriasis further support an inflammatory-multisystem component of meniscus risk^22^. These findings provide genomic evidence for systemic comorbidity clusters in meniscal pathology, motivating a shift toward viewing meniscal degeneration in a broader systemic context

Genome-wide genetic correlation analyses corroborated and extended the PRS-PheWAS findings. Meniscus degeneration showed the strongest overlap with knee osteoarthritis (r_g_ = 0.82–0.89), consistent with its role as both a precursor and accelerator of OA^23-25^. Correlations with obesity and related metabolic traits further reinforce the genetic basis of mechanical and inflammatory contributions to meniscal pathology^11, 26 27^. While most signals reflected cartilage-related biology, modest correlations with inflammatory arthropathies such as rheumatoid arthritis indicate that immune-related pathways may also contribute. Collectively, these results underscore the polygenic architecture shared between meniscus degeneration and systemic comorbid conditions.

GTEx analyses of lead GWAS loci provided biological context for the observed genetic associations. GDF5 and SOX5 were enriched in connective tissue cell types, consistent with their established roles in skeletal development and cartilage integrity^28-30^. BMP6 demonstrated preferential expression in adipose tissue, linking meniscus risk alleles to metabolic pathways relevant to obesity and inflammation^15, 31^. Other loci exhibited broad or testis-specific expressions, suggesting systemic or pleiotropic functions. These patterns indicate that meniscus-associated loci encompass both tissue-specific regulators of cartilage biology and genes with roles in broader metabolic and systemic processes, aligning with the comorbidity landscape revealed by PRS-PheWAS.

Several limitations should be considered. First, the PRS captures only a fraction of genetic liability, and individual effect sizes were modest, underscoring that predictive utility lies in risk pattern identification rather than individual-level prognosis. Second, analyses were conducted in predominantly European ancestry populations, limiting generalizability. Third, PRS associations may reflect indirect pathways; for example, signals such as postoperative infections or complications of orthopedic devices are likely mediated by downstream surgical interventions rather than direct genetic effects. Finally, while PRS and LDSC approaches identify shared genetic architecture, they cannot establish causality. Future studies leveraging Mendelian randomization, diverse ancestries, and functional follow-up of implicated genes will be essential.

Despite these limitations, our study demonstrates that meniscus degeneration is embedded within a systemic genetic architecture linking musculoskeletal, metabolic, and psychosocial domains.

These findings support a paradigm shift in which meniscal pathology is recognized not only as a localized joint disorder but as part of a broader, genetically determined network of comorbidities.

## 4. Conclusions

This phenome-wide analysis shows that genetic risk for meniscus degeneration is embedded in systemic metabolic, inflammatory, and psychosocial pathways rather than being confined to the knee. These insights support integrative management strategies and illustrate how PRS-PheWAS can advance precision medicine in musculoskeletal disease by uncovering shared genetic architectures across health domains

## 5. Methods

### 5.1. Study population

We analyzed data from the UKB, a prospective cohort of approximately 500,000 participants aged 40–69, recruited throughout the United Kingdom between 2006 and 2010^32^. The UKB includes extensive baseline health assessments, genome-wide genotyping, and longitudinal health outcomes obtained via linked hospital inpatient, death, and cancer registries. For this study, we restricted analyses to participants of European ancestry (self-identified White British with consistent genetic ancestry) to minimize population stratification. Individuals with sex chromosome aneuploidy, sex mismatch, or poor genotype quality control were excluded according to the UK Biobank pipeline^33^. All participants provided written informed consent at enrollment, and the study was conducted under UKBB ethical approval (Application Number: 32133)

### 5.2. Genotyping and polygenic risk score

The genotyping procedures and arrays used by the UKB have been described in detail elsewhere^33^. At the sample level, we restricted the analyses to participants who self-identified as White British and exhibited similar genetic ancestry, as determined by principal component analysis of genotypes. Individuals were excluded if they showed discrepancies between self-reported and genetically inferred sex, carried sex chromosome configurations other than XX or XY, or had outlier heterozygosity or missingness rates as defined by the UKB. We further restricted to the high-quality, unrelated subset of samples included in the UKB principal component analysis^33^. At the variant level, we excluded variants with an imputation info score < 0.3 or minor allele frequency (MAF) < 0.01^34^. After quality control, 323,999 samples and 9,505,768 variants remained for analysis.

PRS for meniscus degeneration were constructed using genome-wide summary statistics of meniscus derangement (M13_MENISCUSDERANGEMENTS) from FinnGen Release 12, comprising 32,726 cases and 296,653 controls of Finnish ancestry^35^. We applied PRS-CS, a Bayesian regression framework with continuous shrinkage priors, using the European subset of the 1000 Genomes Project as the external linkage disequilibrium (LD) reference panel^36^. For each UKBB participant, the PRS was computed as the sum of allele dosages weighted by the posterior effect sizes estimated by PRS-CS. PRSs were standardized to z-scores with a mean of 0 and a standard deviation of 1 prior to analysis.

### 5.3. Phenotype definitions

We conducted PheWAS of the meniscus PRS against a broad array of disease phenotypes derived from UKB health records. Phenotypes were defined using the established PheCODE 1.2 mapping system, which aggregates related ICD-9 and ICD-10 diagnosis codes into clinically meaningful case definitions (e.g., grouping all ICD codes for type 2 diabetes mellitus under a single phenotype)^37^. Case status required at least one instance of the diagnosis code in hospital inpatient or self-reported medical records, and control status was defined as the absence of the code. An exclude range for each phenotype was applied to filter out controls with related conditions, as implemented in the PheWAS R package^38^. To ensure reliable association estimates and avoid unstable results, we excluded phenotypes with extremely low case counts. Phenotypes with fewer than 200 cases or controls were excluded, resulting in 962 phenotypes retained for downstream analyses. Key broad phenotype groupings included endocrine/metabolic disorders, mental health disorders, neurological diseases, cardiovascular conditions, respiratory diseases, gastrointestinal/hepatobiliary disorders, musculoskeletal disorders, dermatologic/immunological diseases, and injury/poisoning categories, among others, aligning with the established phenotype classification schema. The target trait of interest, meniscus degeneration, was defined using ICD-9 codes 717.0–717.5 and ICD-10 codes M23.0–M23.3, including all subcodes.

### 5.4. Polygenic risk score-based phenome-wide association study

For each phenotype, logistic regression was performed, adjusting for demographic and genetic covariates using the following formula:

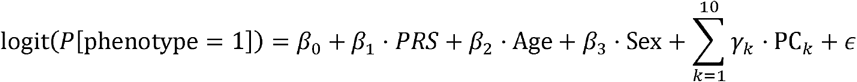

Where *P* [phenotype = 1] is the probability of having the phenotype, PRS is the individual’s polygenic risk score for meniscus degeneration, and PC_k_ are the first 10 genetic principal components are used to account for population structure. P-values were calculated using Wald statistics and adjusted for multiple comparisons using the Bonferroni correction (α = 5.20×10^-5^, 0.05/962).

### 5.5 Sensitivity analysis

To assess the robustness of our findings and disentangle overlapping genetic risk of the meniscus degeneration PRS from the direct consequences of a clinical meniscus degeneration diagnosis, we conducted two complementary sensitivity analyses. First, we repeated the primary PheWAS while additionally adjusting for meniscus degeneration status as a covariate, thereby testing whether associations persisted after accounting for an individual’s clinical diagnosis. Second, we excluded all individuals with a diagnosis of meniscus degeneration and re-ran the PheWAS in the remaining disease-free subset, allowing us to assess whether the observed associations were independent of case status.

We additionally conducted stratified PheWAS analyses to evaluate potential effect modification by demographic and lifestyle factors. The cohort was stratified by sex, smoking status (ever vs. never), and BMI categories based on clinical cutoffs. Within each subgroup, we repeated the PheWAS, modeling the association between the meniscus degeneration PRS and each phenotype to assess the consistency of associations across subgroups.

### 5.6. Linkage Disequilibrium Score Regression

We estimated genetic correlations between meniscus degeneration and other phenotypes using linkage disequilibrium score regression (LDSC)^39^. Publicly available GWAS summary statistics from FinnGen release 12 were used for all phenotypes, including the target trait M13_MENISCUSDERANGEMENTS. Summary statistics were filtered to HapMap3 variants and restricted to well-imputed SNPs (INFO > 0.9) with minor allele frequency ≥ 0.01. LDSC was performed with precomputed LD scores from the European population of the 1000 Genomes Project reference panel^40^. Genetic correlation (r_g_) estimates were obtained for each phenotype pair, and standard errors were computed using block jackknife resampling as implemented in LDSC. Phenotypes included in the analysis were those identified as significantly associated with the meniscus degeneration PRS in the PRS-PheWAS meta-analysis.

### 5.7. GTEx functional annotation

We evaluated the tissue-specific expression of lead genes identified from the FinnGen meniscus degeneration GWAS using the Genotype-Tissue Expression (GTEx) Project, version 10^41, 42^. Median transcript abundance values (transcripts per million, TPM) across 68 tissues were obtained from the GTEx v10 reference dataset. For each gene, we computed the tissue-specificity index (τ), which ranges from 0 (ubiquitous expression) to 1 (tissue-restricted expression), to quantify tissue selectivity^43^. The top tissue and its corresponding median TPM were reported for each gene. Genes were subsequently categorized as tissue-specific (τ ≥ 0.85) or broadly expressed (τ < 0.60), following prior studies.

## Data Availability

All data produced in the present study are available upon reasonable request to the authors.

## Acknowledgements

This work was supported by NIGMS R01 GM138597. We acknowledge the participants and investigators of the FinnGen study, and all the participants of the UK Biobank. The use of the UK Biobank resources was approved under Application Number 32133.

## Notes

### Competing Interest Statement

The authors have declared no competing interest.

